# Mifepristone as a pharmacological intervention for stress-Induced alcohol craving: a translational crossover randomized trial

**DOI:** 10.1101/2023.01.02.23284122

**Authors:** Carolina L. Haass-Koffler, Molly Magill, Nazzareno Cannella, Joshua C. Brown, Elie G. Aoun, Patricia A. Cioe, Rajita Sinha, Robert M. Swift, Roberto Ciccocioppo, Lorenzo Leggio

## Abstract

Preclinical and clinical work suggests that mifepristone (glucocorticoid receptor antagonist), may be a viable treatment for alcohol use disorder (AUD). The aim of this work was to translate our preclinical mifepristone study using yohimbine (α2 receptor antagonist) stress-induced reinstatement of alcohol-seeking to a clinical setting. This was a Phase 1/2, outpatient, cross-over, randomized, double-blind, placebo-controlled trial with non-treatment-seeking individuals with AUD (*N*=32). We investigated the safety, alcohol craving and consumption after oral administration of mifepristone (600mg daily for a week) in a human laboratory study comprised of administration of yohimbine in a cue-reactivity procedure and alcohol self-administration. Outcomes were assessed using Generalized Estimating Equations and mediation and moderation analyses assessed mechanisms of action and precision medicine targets. We did not observe serious adverse events related to the study drugs or study procedure and mild to moderate non-serious adverse events were reported by both study conditions. Also, there was no statistically-significant difference between the mifepristone and placebo in the hemodynamic response, alcohol subjective effects and pharmacokinetics parameters. Mifepristone significantly reduced alcohol craving and increased cortisol levels. Mifepristone-induced cortisol increase was not a mediator of alcohol craving. Moderation analysis with family history density of AUD (FHDA) and mifepristone, suggested that reduced craving was present in individuals with *low*, but not *high* FHDA. Mifepristone, compared to placebo, did not reduce alcohol consumption in the laboratory or in a naturalistic setting. This study successfully translated a preclinical paradigm to a human laboratory study confirming safety, tolerability and efficacy of mifepristone in an alcohol paradigm. Mediation analysis showed that the effect of mifepristone on craving was not related to mifepristone-induced increases in cortisol and moderation of FHDA suggested the importance of evaluating AUD endophenotypes for pharmacotherapies.

**Clinical trial registration:** Clinicaltrials.gov; NCT02243709

**IND/FDA:** 121984, mifepristone and yohimbine (Holder: Haass-Koffler)

## Introduction

Stress on biological systems has been linked to depression, anxiety^1^ and alcohol use disorder (AUD)^2,3^. Stress combined with re-exposure to priming or to environment and cues previously associated with alcohol exacerbates reoccurring drinking episodes both in rodents^4-6^ and humans^7,8^.

Mifepristone, a glucocorticoid receptor antagonist, is a medication approved by the FDA for the treatment of hyperglycemia, secondary to endogenous Cushing syndrome, in adults who have failed surgery or are not candidates for surgery, as well as for the termination of early pregnancy. Mifepristone has been under investigation as a potential treatment for many neuropsychiatric disorders such as psychotic depression (data pooled from three studies: mifepristone *n*=833; placebo *n*=627)^9^, including AUD (mifepristone *n*=28; placebo *n*=28)^10^.

In our preclinical work, we demonstrated that systemic administration of mifepristone, as well as its infusion in the central nucleus of the amygdala, reduced yohimbine-induced reinstatement of alcohol-seeking in alcohol-dependent Long Evans rats^11^. The effect of mifepristone on reducing alcohol-seeking was also supported by studies in alcohol-dependent Wistar rats^10^.

One of the most challenging aspects in designing a human laboratory study is the inclusion of an acute stress condition intended to represent a comprehensive naturalistic environment in individuals with AUD. In this study, we utilize yohimbine, rather than other stressors (psychological, physical), to maximize the translational efforts from our preclinical model^11^. Yohimbine is a well-validated pharmacological tool^12^ that has been widely employed in preclinical alcohol research studies to evaluate the effect of noradrenergic activation as a proxy for physiologic stress^11,13,14^. As a pharmacological challenge, yohimbine was shown to activate the hypothalamic-pituitary-adrenal (HPA) axis, in addition to increasing sympathetic nervous system activity^15^ and increasing alcohol craving^16^. This model builds on the evidence that the glucocorticoid receptor plays a role in the reinstatement behaviors elicited by yohimbine^11^, and that noradrenergic activation has been linked to alcohol intake and reoccurrence during abstinence^17^.

Translational studies should evaluate discrepancies between preclinical and clinical studies to reduce the knowledge gap between preclinical and clinical setting. In a study of a rhesus macaque AUD model^18^, cortisol was reported as potential mediator of mifepristone effect on alcohol self-administration whereas in a trial with pooled clinical studies, mifepristone plasma concentration was a more powerful mediator of mifepristone effects on psychiatric symptoms than cortisol^9^. It is also important to evaluate possible discrepancies between different preclinical models of AUD to better evaluate precision medicine approaches based on AUD endophenotype. Mifepristone reduced alcohol self-administration in Wistar rats trained to consume alcohol, however, marchigian sardinian alcohol preferring (msP) rats, a genetically selected AUD model, were less responsive to mifepristone’s ability to reduce alcohol self-administration^19^. This information when translated in human laboratory study may inform pharmacotherapy approached based on family contribution to the development of AUD^20^.

The aim of this work was to translate our preclinical study on the effect of mifepristone on yohimbine stress-induced reinstatement of alcohol-seeking^11^ in a human laboratory study. The primary outcome of this study was to test in individuals diagnosed with AUD, the safety of oral administration of mifepristone (600mg/day/7-day), compared to placebo. We used a human laboratory paradigm comprised of oral administration of yohimbine (32.4mg/1-day) paired to a cue-reactivity procedure, a priming alcohol dose and alcohol self-administration in an open bar laboratory. Secondary outcomes included: assessment of alcohol craving and consumption during the laboratory procedures. Finally, mediation and moderation analyses were conducted to assess potential mechanisms of action and precision medicine targets.

## Materials and methods

### Study design, setting and approval

The study was a Phase 1/2, outpatient, cross-over, randomized, double-blind, placebo-controlled, human laboratory study (**Figure 1**) and reported following the Consolidated Standards of Reporting Trials (CONSORT) extension^21^. A crossover design was chosen for this study because the within-subject variation is less than the between subject variation and allows for recruitment of less participants. The study was conducted at the Center for Alcohol and Addiction Studies, Brown University, Providence, RI, USA from 2014-2021. The trial was approved by the Brown University Institutional Review Board, conducted under an FDA Investigational New Drug application (IND121984) and registered at clinicaltrials.gov (NCT02243709). Important amendment to the original clinical study included the inclusion of females within childbearing age, using non-hormonal, barrier contraceptive, to improve female representation in the study (February-1, 2019).

**Figure 1.**
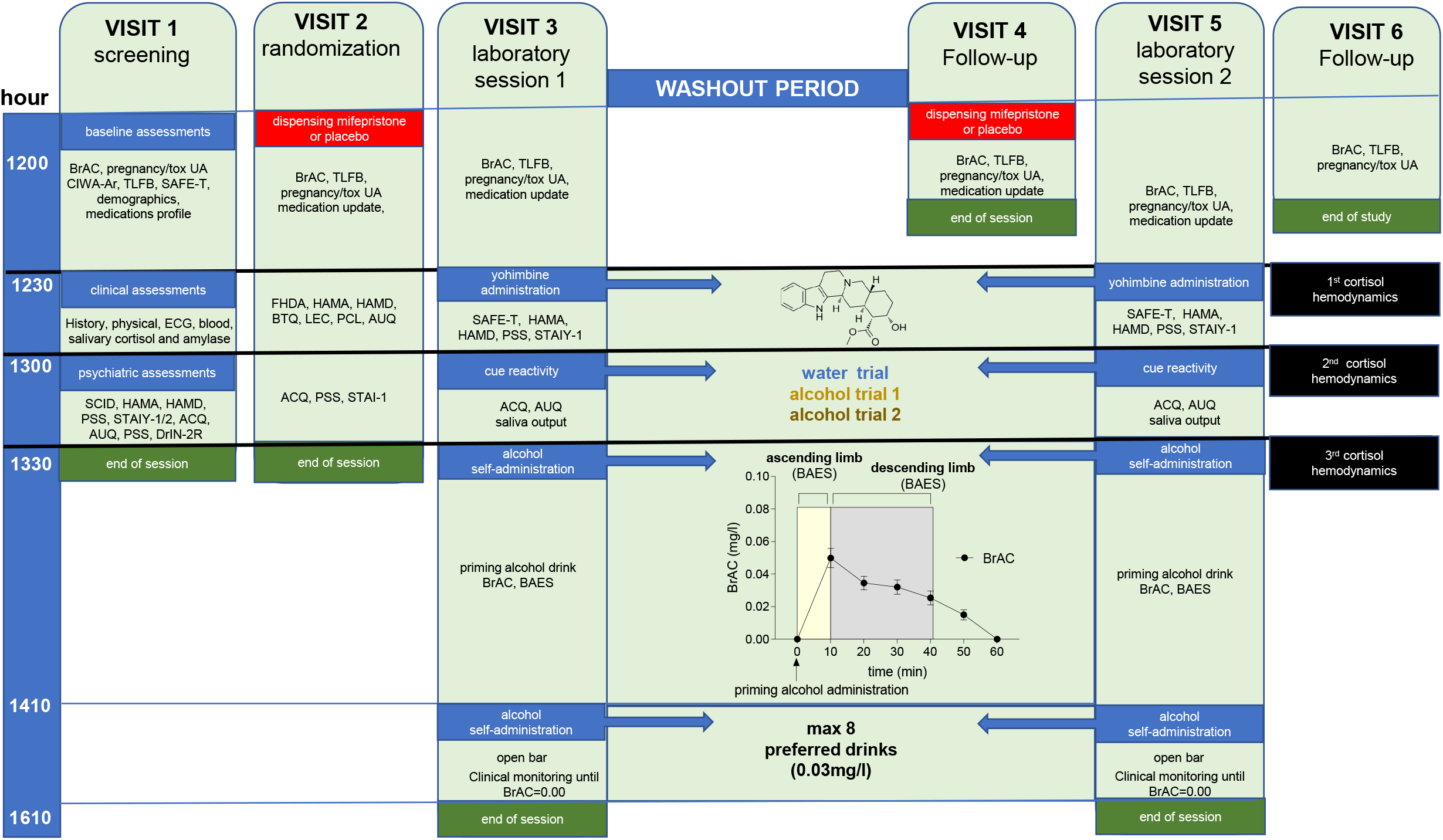
Schematic outline of the laboratory paradigm procedures. Visit 1 (screening), Visit 2 (randomization, mifepristone or placebo), Visit 3: laboratory 1 (mifepristone or placebo), washout period (21 days), Visit 4 (follow-up and second condition: placebo or mifepristone) and Visit 5: laboratory 2 (opposite condition, counter balanced), Visit 6 (follow up). *Legend*: ADS, Alcohol Dependence Scale; ACQ, Alcohol Craving Questionnaire; AUQ, Alcohol Urge Questionnaire; BAES, Biphasic Alcohol Effects Scale; BrAC, breath alcohol concentration; BTQ: Brief Trauma Questionnaire; CIWA-Ar, Clinical Institute Withdrawal Assessment for Alcohol-revised; ECG, electrocardiogram; FHDA, Family History Density of Alcoholism; HAMA/HAMD, Hamilton Anxiety and Depression Rating Scale; LEC, Life Event Checklist; PLC, PTSD Checklist; PSS, Perceived Stress Scale; SAFE-T, Suicide Assessment Five-Step Evaluation and Triage; SCID, Structured Clinical Interview for DSM-IV, STAI, Spielberger State Trait Anxiety; TLFB, Timeline Followback; UA, urine analysis

### Participants

After signing a written informed consent document and performing a screening to assess inclusion/exclusion criteria (**Methods S1**), eligible, non-abstinent, individuals with AUD who were no seeking treatment, were randomly assigned, by computer allocation, to 7-day treatment with either daily 600mg mifepristone or placebo. After a 3-week washout period to allow cortisol levels to return to baseline after mifepristone administration^22^ and avoiding carryover effect, participants returned to the laboratory and received the crossover condition.

### Study procedures, drugs, dose justification and compliance

Details of visit 1 (*screening*), visit 2 (*randomization*), visit 3 and 5 (*laboratory sessions*) and visit 4 and 6 (*follow up*) are published^23^ and reported in **Methods S1**. Mifepristone and matching placebo were provided by Corcept Therapeutics (Menlo Park, CA). The oral dose of yohimbine was based on studies that examined neuroendocrine activation in humans^16,24,25^ and compounded by a local pharmacy.

### Statistical analysis

For all outcomes, we utilized an intention-to-treat (ITT) approach, where participants were examined based on their *a priori* randomized protocol and received at least one dose of the study medication (mifepristone or placebo)^26^. All analyses were conducted using Generalized Estimating Equations (GEE)^27^ with robust standard errors, and an unstructured correlation matrix, unless noted.

#### Primary outcomes

safety and tolerability assessed by the number of adverse events (AEs) of oral administration of mifepristone was assessed after 7 days in an outpatient setting, and when it was administered with yohimbine and alcohol during the laboratory paradigms. Additional safety and tolerability analysis included hemodynamic response, alcohol pharmacokinetics and subjective effects of alcohol between the mifepristone and placebo condition (*χ*^*2*^ test).

#### Secondary outcomes

Craving measures included alcohol craving questionnaire short form-revised (ACQ-SF-R)^28^. Additional craving analysis included alcohol urge questionnaire (AUQ)^29^, and cue-induced saliva output. Values of the water trials for each dependent variable were inserted as covariate in the model (allowed for the dependent variable to be specific for alcohol), time coded: t_1_=alcohol trial 1 and t_2_=alcohol trial 2. In the bar laboratory, alcohol consumption was measured by number of drinks consumed (*t*-test). In the outpatient setting, alcohol consumption was measured by self-report using the timeline follow back method, reported as heavy drinking days and drinks per week.

#### Mediation

analyses for cortisol level, on ACQ, AUQ and cue-induced salivary output, were conducted using a regression-based, Macro Estimating Model^30^ that estimated the indirect effect of a within-participant manipulation on outcomes. Mediation was tested using standard procedures (product of the *a* and *b* path coefficients), but difference scores were created for the mediator and outcome under mifepristone/placebo conditions. The dependent variables were ACQ, AUQ and cue-elicited salivary output and the mediator was the cortisol level after 7-day mifepristone (M_1_) or placebo (M_2_) administration. The indirect effect was tested with Monte Carlo *CI* (interval estimate confidence 95%).

#### Moderator

analyses for family history density of alcoholism (FHDA) on ACQ, AUQ and salivary output were conducted using dichotomous predictor variables^31^. Consistent with previous work^32^, we used the Family Tree Questionnaire^33^ to calculate the FHDA and dichotomized (median split into *low/high*)^34^. FHDA was set as the moderator (m_0_=FHDA *low*; m_1_=FHDA *high*) and drug (mifepristone/placebo) as within-subject factors. For the outcomes in the laboratory (ACQ, AUQ, cue-elicited salivary output and neuroendocrine variations), the model was specified to evaluate: main effect of FHDA, drug, and the moderator by drug interaction. For additional statistical details, see **Method S1**.

## Results

### Participants’ characteristics

The CONSORT diagram is reported in **Figure 2** and sociodemographic and baseline clinical characteristics of the participants in **Table S1**. One hundred-fifty-five participants were screened on the telephone, 46 were screened in person and 32 were randomized. They received at least one dose of the study medication and included in ITT analysis. Additional information on participant retention can be found in the **Results S1**.

**Figure 2.**
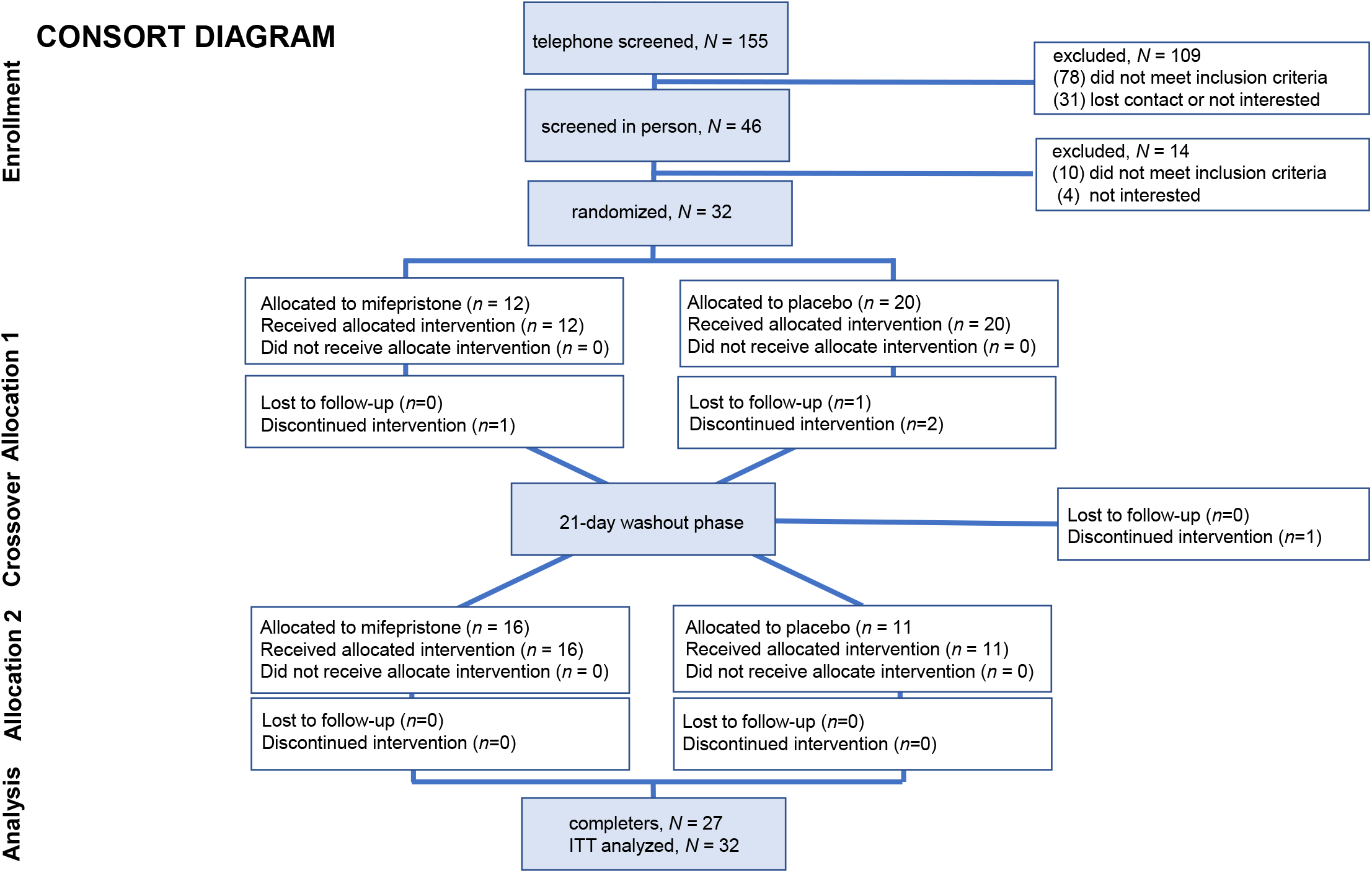
Consolidated Standards of Reporting Trials (CONSORT) extension for cross-over trials.

### Primary outcome

#### Safety and tolerability of mifepristone, yohimbine and alcohol

There were no serious AEs when the study medication was co-administered with yohimbine and alcohol in the laboratory. We observed three non-serious AEs (mifepristone: *n*=0, 0%; placebo: *n*=3, 10%; *p*>.05). Two individuals had an emesis episode after yohimbine and alcohol administration and one individual experienced increased blood pressure after yohimbine administration but before alcohol administration, however, blood pressure normalized after the priming alcohol administration.

The safety and the tolerability of the laboratory procedures were also assessed by monitoring the hemodynamic function (SBP, DBP and HR), alcohol pharmacokinetic parameters (C_max_, T_max_ and AUC_0-40_) and subjective response to alcohol (stimulation/sedation) (**Figure 3**). For SBP, we found no significant main effect for drug, a significant time effect such that SBP increased from baseline after cue-reactivity (*p*<.013), and a time by drug interaction, where these increases were observed only in the placebo after the cue-reactivity (t_60min_, *p*=.020) (**Figure 3A**). For DBP, we found a significant main effect for drug, such that DBP was lower in the mifepristone condition compared to placebo (*p*=.005), a main effect for time such that DBP increased from baseline to before (t_30min_, *p*=.001) and after (t_60min_, *p*=.002) the cue-reactivity, and a time by drug interaction, where these increases were observed only in the placebo after the cue-reactivity (t_60min_, *p*<.001) (**Figure 3B**). Finally, for HR, there was no significant main effect for drug, time or drug by time interaction (*p’s*>.05) (**Figure 3C**).

**Figure 3.**
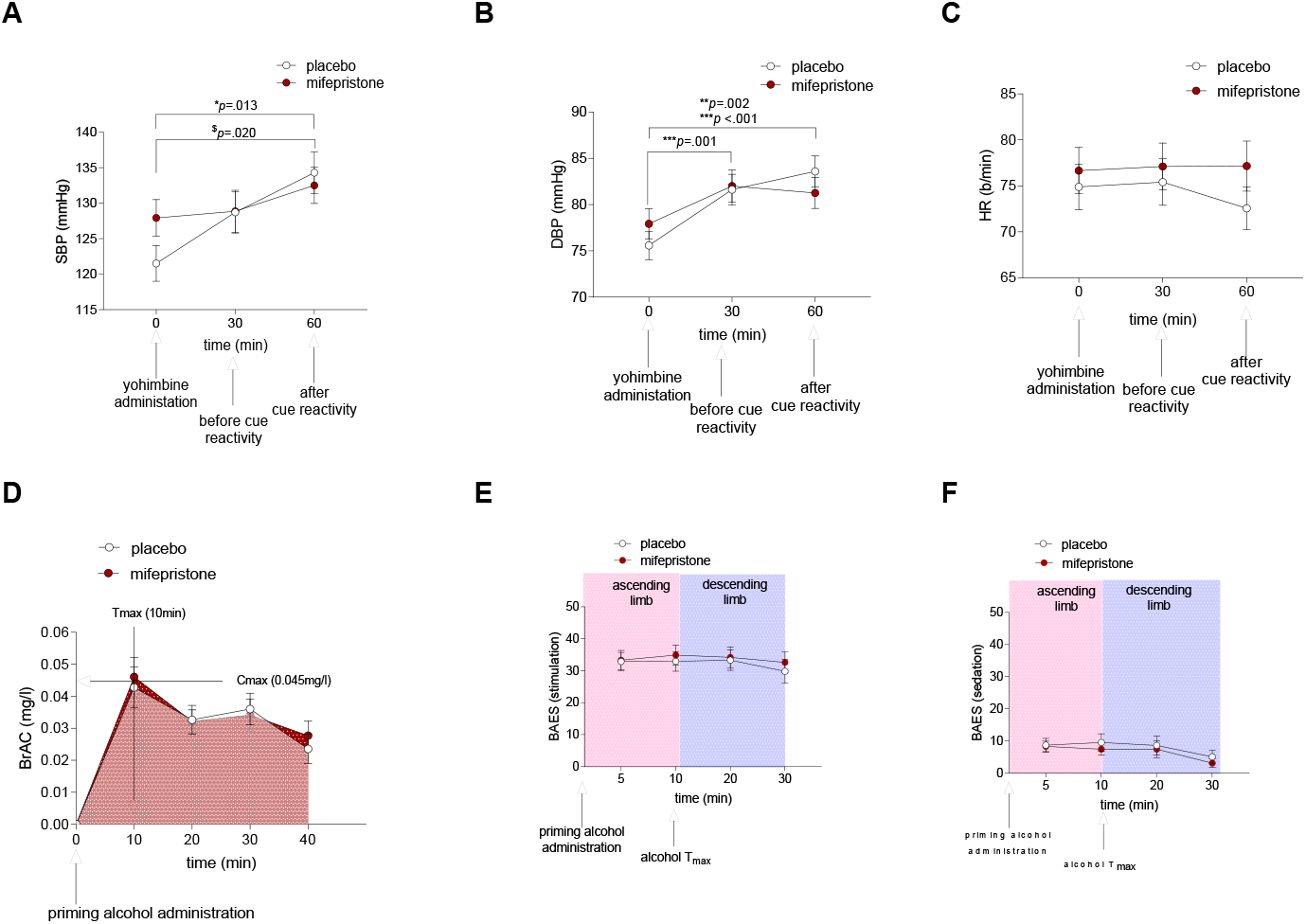
Hemodynamic function, subjective response and pharmacokinetic parameters after administration of yohimbine paired to a cue reactivity and alcohol self-administration paradigm. **A**) SBP: no significant main effect for medication, a significant time effect such that SBP increased from baseline to after cue-reactivity (t_60min_, *b=*5.96; *CI*=1.27; 10.65; *p*<.013), and a time by medication interaction, where these increases were observed only in the placebo condition after the cue-reactivity (t_60min_, *b=*8.149; *CI=*1.27, 15.03, *p*=.020). **B**) DBP, a significant main effect for medication, such that DBP was lower in the mifepristone condition compared to placebo (*b=-*4.01; *CI*=-6.80, -1.215; *p*=.005), a main effect for time such that DBP increased from baseline to before (t_30min_, *b=*3.639; *CI=*1.41, 10.21; *p*=.001) and after (t_60min_, *b=*4.79; *CI*=1.83, 7.75, *p*=.002) the cue-reactivity, and a time by medication interaction, where these increases were observed only in the placebo condition after the cue-reactivity (t_60min_, *b=*6.221; *CI*=2.57, 9.88, *p*<.001). **C**) HR: no significant main effect for medication, time or medication by time interaction (*p’s*>.05). All data presented as mean±*SEM*. ^*^*p*<.05 main effect; ^$^*p<*.05 interaction.

After the yohimbine administration and cue-reactivity procedure, to assess the safety and tolerability of mifepristone when co-administered with alcohol, we measured alcohol pharmacokinetics via BrAC after the administration of a prime alcohol drink designed to rise BrAC to 0.03-0.05mg/l. There was no difference in mifepristone condition compared to placebo in the BrAC pharmacokinetic curve parameters (AUC, T_max_, C_max_) (*p’s*>05) (**Figure 3D**). Also, there was no significant difference in the mifepristone compared to placebo (main effect and interaction), on the alcohol subjective effect both in stimulation and sedation (*p’s*>.05) scales (**Figure 3E-F**). During the 7-day administration of mifepristone or placebo in an outpatient setting, with mifepristone, we did not observe serious AEs related to the study drugs or study procedure (**Table S2, Results S1**). Mild to moderate non-serious AEs were reported by both study conditions throughout the trial, with no difference (*p’s*>.05).

### Secondary outcomes

#### Alcohol craving, urge and cue-elicited salivary output

After the administration of yohimbine, the effect of mifepristone, compared to placebo, on alcohol craving was assessed during a cue-reactivity procedure using ACQ. Additional analysis included AUQ and cue-elicited salivary response. Analysis of ACQ showed no main effect for drug (*p*>.05), but a significant main effect for time (*p<*.001) where increases of craving were observed in alcohol trial 2. A time by drug interaction suggested decrease of craving for the mifepristone compared to placebo (*p=*.007) at the alcohol trial 1 (**Figure 4A**). Analysis of AUQ showed a main effect for drug (*p=*.010) where a decrease in urge was observed in mifepristone compared to placebo, but not a main effect for time (*p*>.05). We found, however, a drug by time interaction in the alcohol trial 1 (*p=*.040), suggesting a lower urge in mifepristone compared to placebo. (**Figure 4B**). Analysis of cue-elicited salivary output revealed a significant main effect for drug (*p<*.001), where lower saliva output was observed in the mifepristone compared to placebo, and a significant main effect for time showed salivary decreases at the alcohol trial 2 (*p*<.001). A time by drug interaction was observed both at the alcohol trial 1 (*p<*.001) and alcohol trial 2 (*p<*.001), with decrease of salivary output in the mifepristone condition compared to placebo (**Figure 4C**).

**Figure 4.**
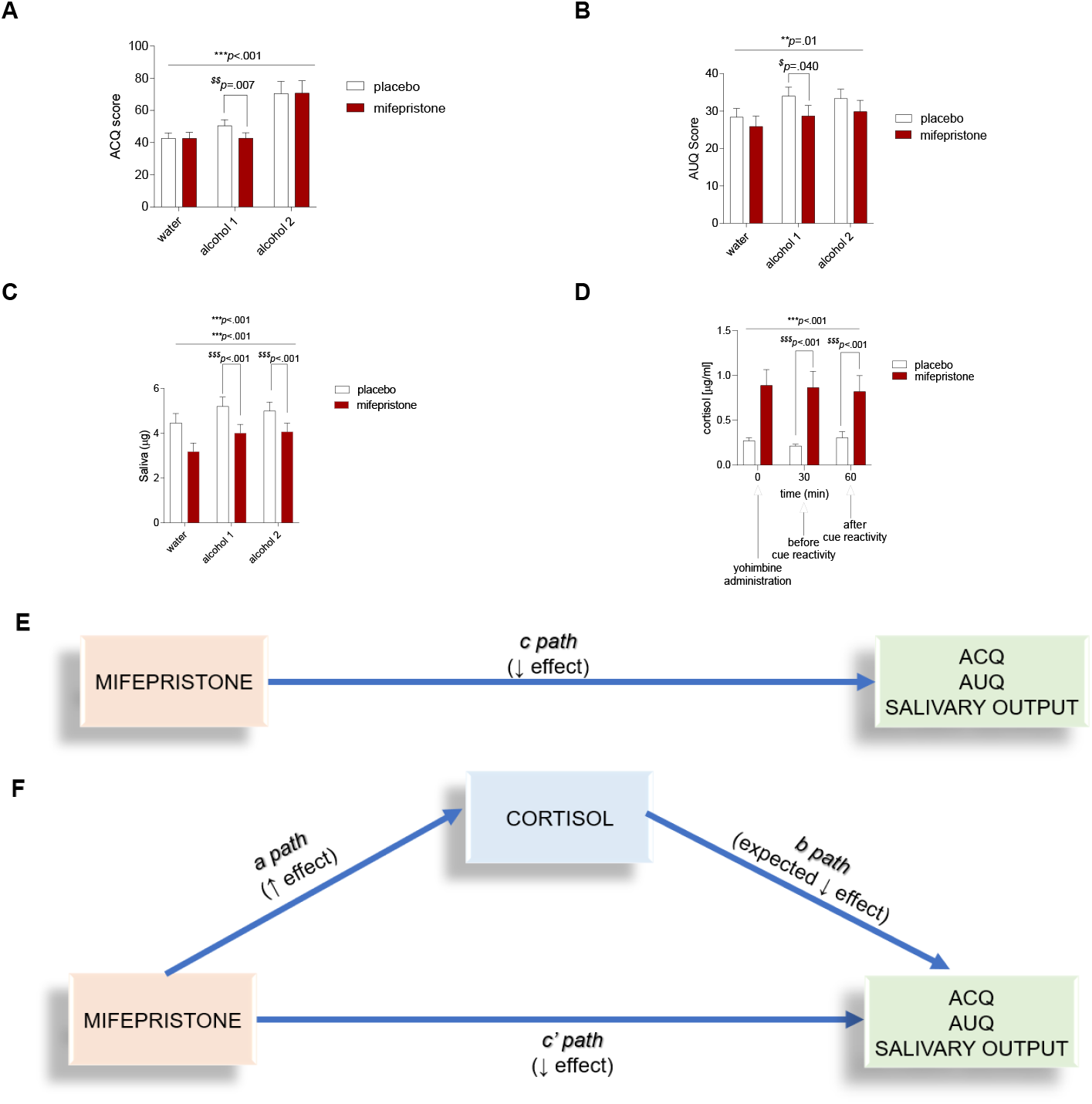
The effect of mifepristone compared to placebo on alcohol craving, urge and cue-elicited salivary output and cortisol and mediation modle. **A**) ACQ: no main effect for medication (*p* >.05), but a main effect for time (*p<*.001) and a time by medication interaction (*p=*.007) at the alcohol trial 1. **B**) AUQ: a main effect for medication (*p=*.010), no main effect for time (*p*>.05), but a medication by time interaction in the alcohol trial 1 (*p=*.040). **C**) cue-elicited salivary output: a main effect for medication (*p<*.001), a main effect for time (*p*<.001) at the alcohol trial 2, and a time by medication interaction at the alcohol trial 1 (*p<*.001), and alcohol trial 2 (*p<*.001). **D**) cortisol: a significant main effect for medication (*b*=.54; *CI*=.22, .85; *p*<.001), no main effect for time (*p*>.05), but there was a medication by time interaction both before (t_30min_: *b=*.634; CI=.31, .99: *p*<.001) and after (t_60min_: *b=*.49; *CI*=.29, -.91 *p*<.001) the cue reactivity. *Mediation Model*. Increase of cortisol level as mediator of alcohol craving, urge and salivary output after 7-day mifepristone administration before initiating any laboratory procedure. **E**) Total effect (*c path*); **F**) Direct effect (*c’ path*); Indirect effect (*a x b path*). ↓↑ represent effect of mifepristone on outcomes. All data presented as mean±*SEM*. ^*^*p*<.05 main effect; ^$^*p<*.05 interaction.

#### Cortisol

Analysis of saliva cortisol during the laboratory procedures, revealed a significant main effect for drug (*p*<.001), such that higher cortisol was observed in the mifepristone, compared to placebo, no main effect for time (*p*>.05), but there was a drug by time interaction, indicating higher cortisol levels both before (t_30min_ *p*<.001) and after (t_60min_ *p*<.001) the cue-reactivity (**Figure 4D**). This result further supports individuals’ medication compliance as cortisol increases with mifepristone administration^22^.

Finally, to test if participants responded to the laboratory procedures, the increase of the HPA was confirmed when we compared the value of cortisol levels collected at the screening visit (basal) to the values collected during the laboratory visits (stress) in the placebo (**Results S1, Figure S1**).

### Cortisol as mediator of alcohol craving, urge and cue-elicited salivary output

The above results support the total effect of mifepristone on ACQ, AUQ, and cue-elicited salivary output outcomes at alcohol trial 1 (*c path*) (**Figure 4E**), as well as an effect of mifepristone on cortisol level (*a path*, described above **Figure 4D**). However, *b path* results did not show a relationship between cortisol and ACQ (*p*>.05), AUQ (*p*>.05) and cue-elicited salivary output (*p*>.05) at the alcohol trial 1. We also tested the effect combining alcohol 1 and 2: ACQ (*p*>.05), AUQ (*p*>.05) and cue-elicited salivary output (*p*>.05). As a result, the Monte Carlo *CI* around the product of the *a* and *b path* coefficients were non-significant (*p’s*>.05) (**Figure 4F**).

### FHDA as moderator of alcohol craving, urge, cue-elicited salivary output and cortisol

For ACQ, there was a main effect for FHDA (*p*=.023) such that craving was higher for those with *high* FHDA compared to *low* FHDA. There was also a FHDA by drug interaction, suggesting lower craving in the mifepristone, compared to placebo, in those with *low* FHDA (*p*=.007) (**Figure 5A**). Similarly, for AUQ, there was a main effect for FHDA (*p*=.018), with increased urge in the *high* FHDA compared to the *low* FHDA group, and a FHDA by drug interaction (*p*=.028), suggesting lower urge in the *low* FHDA group for mifepristone, compared to placebo condition (**Figure 5B**). For cue-elicited salivary output, we found a main effect for FHDA (*p*=.035) with increased salivation in the *high* FHDA compared to the *low* FHDA group. In the FHDA by drug interaction analysis, we found that mifepristone, compared to placebo, reduced salivary output both in the *low* FHDA (*p*=.025) and in the *high* FHDA (*p*=.002) groups with greater reductions for *high* FHDA participants (**Figure 5C**). When we tested if FHDA was a moderator of HPA-axis response, we found a main effect of FHDA (*p=*.049), such that lower cortisol was observed in the FHDA *high* compared to the FHDA *low* participants. There was also a FHDA by drug interaction, such that there was higher cortisol level both in the FHDA *low* (*p=*.042) and in the FHDA *high* (*p=*.036) groups with greater increases for high FHDA participants (**Figure 5D**).

**Figure 5.**
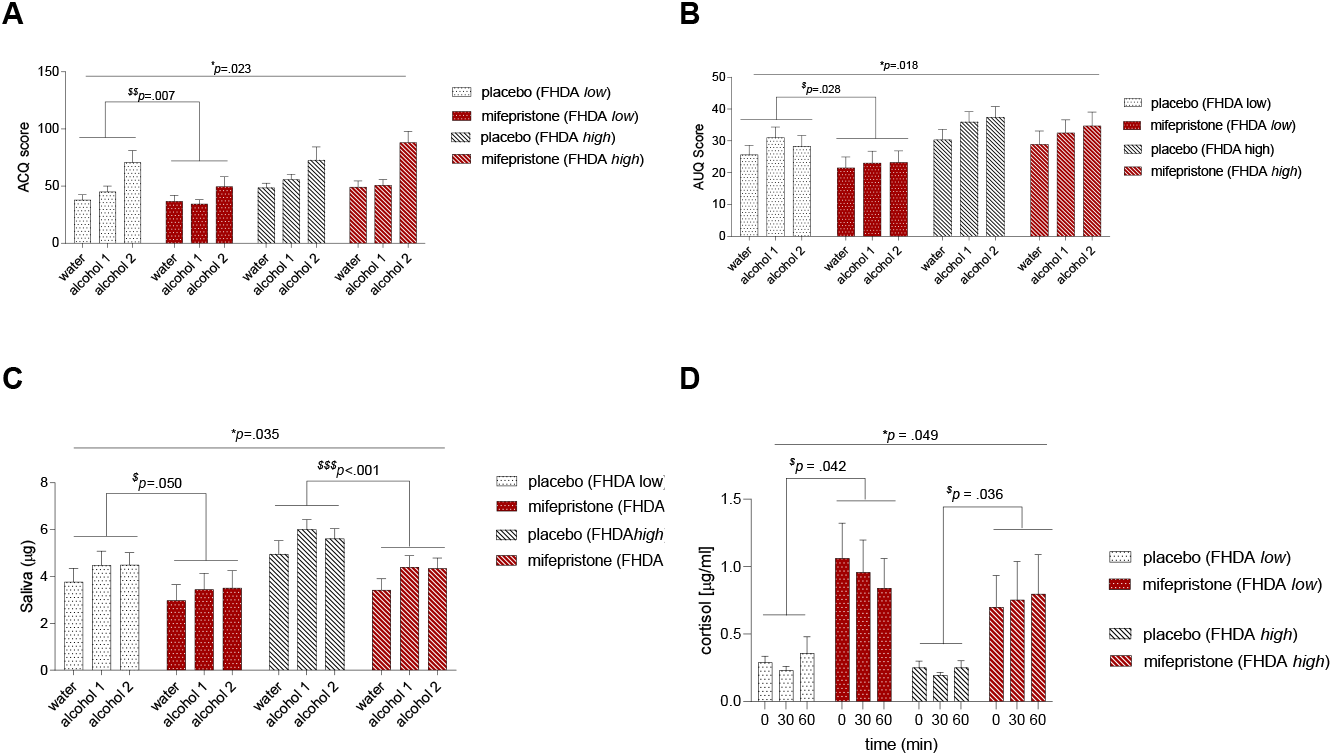
FHDA moderation analysis: the effect of mifepristone compared to placebo on alcohol craving, urge and cue-elicited salivary output. **A**) ACQ: a main effect for FHDA (*b*=17.36; *CI=*2.43, 32.29; *p*=.023), a FHDA by medication interaction in those with *low* FHDA (*b*=-18.65; *CI*=-32.10, -5.20; *p*=.007). **B**) AUQ: a main effect for FHDA (*b*=-11.096; *CI*=1.89, 20.30; *p*=.018) in the *high* FHDA compared to the *low* FHDA, a FHDA by medication interaction (*b*=-9.781; *CI*=-18.53, -1.03; *p*=.028) in the *low* FHDA in the mifepristone group, compared to placebo. **C**) Salivary output: a main effect for FHDA (*b*=1.623; *CI*=.12, 3.13; *p*=.035) in the *high* FHDA compared to the *low* FHDA, a FHDA by medication interaction both in the *low* FHDA (*b*=-.588; *CI*=-1.17, -.20; *p*=.025) and in the *high* FHDA (*b*=-2.122; *CI*=-.43, -3.22, *p*=.002). **D**) Cortisol: a main effect of FHDA (*b*=-0.328; *CI*=-32.34, -5.22; *p=*.049), a FHDA by medication interaction, both in the FHDA *low* (*b*=0.413; *CI*=2.10, 5.20; *p=*.042) and in the FHDA *high* (*b*=.708; *CI*=2.15, 5.43, *p=*.036). All data presented as mean±*SEM*. ^*^*p*<.05 main effect; ^$^*p <*.05 interaction.

Participants responded to stress cue-reactivity paradigm (**Results S1, Figure S2**), FHDA was not related to differences in age, age of onset of AUD, alcohol dependence, baseline of craving (ACQ), urge (AUQ), stress (PSS, STAI trait and state), anxiety (HAMA) (all *VIF*<5)^35^ or trauma (assessed by the BTQ and LEC, criterion A for PTSD) (*Spearman, p*>.05), therefore indicating that FHDA was likely an independent moderator.

### Alcohol consumption

In the open bar laboratory session, participants consumed a small number of standard alcohol drinks both in the mifepristone (0.8±0.3) and placebo (0.5±0.2) conditions, with no significant difference between conditions (*p>*.05). In an outpatient setting, neither mifepristone or placebo reduced alcohol consumption (*p’s*>.05) (**Results S1, Figure S3**).

## DISCUSSION

The major finding of this study is that mifepristone, administered with yohimbine and alcohol, was safe for individuals diagnosed with AUD. We also demonstrated that mifepristone, compared to placebo, attenuates combined yohimbine/cue-induced alcohol craving in the laboratory. Mifepristone effect in reducing yohimbine-induced alcohol craving is independent of the mifepristone-induced increase of cortisol level and significantly reduced craving only in individuals with *low* family history of AUD (FHDA).

At the research level, our results cast important relevance as this was the first time that a preclinical paradigm that included the use of a noradrenergic pharmacological stressor (yohimbine)^12^, paired to a cue-reactivity (cue-reinstatement) and alcohol self-administration, was successfully and safely translated in a human laboratory study in individuals with diagnosis of AUD. We effectively translated an extensively used preclinical paradigm to a human laboratory study that investigates stress-induced alcohol craving, including a comprehensive model capable of integrating contributions of each alcohol-proximal variable (e.g., neuroendocrine variations, family history, stress and trauma factors) that may influence craving^36^. The robustness of the laboratory procedures was supported by the increase of blood pressure from baseline post cue-reactivity. The analysis of the laboratory session from baseline (screening) demonstrated that the participants responded to the procedures, as it is known that both cue-reactivity^37^ and yohimbine^24^ increase blood pressure.

At the clinical level, it was critical to assess the safety and tolerability of study drug when co-administered with alcohol, given the potentially serious consequences of drug-alcohol interactions. This assessment is important for novel^38^ and repurposed^39^ medication for treating AUD and consistent with the FDA^40^ and the European Community (EMA/CHMP/EWP/20097/2008) guidelines on the development of medications to treat AUD. Clinical screening included laboratory analysis to evaluate possible overlapping enzymatic pathways and limited renal activity for excretion of multiple drugs. Under this paradigm, we did not encounter serious AEs in either the mifepristone or in the placebo, and non-serious AEs were encountered in both conditions. After the alcohol prime, mifepristone compared to placebo did not did not alter the alcohol pharmacokinetics and did not affect the stimulation/sedation effects of alcohol, supporting the safety of mifepristone when co-administered with alcohol.

Mifepristone reduced the self-reported alcohol craving (ACQ and AUQ) at the first alcohol cue exposure. It is known that the second alcohol trial boosts alcohol craving^41-43^, in this study it is possible that the effect of mifepristone was washed out under the strong stress procedures. The cue-elicited salivary output, however provided an objective biomarker for the effect of mifepristone in craving reduction, in both trials. This observation is in line with original cue-reactivity studies showing that salivation, rather than be associated to conscious attention to alcohol (vision, smell and tactile experience), it is more pronounced in individuals with serious AUD and is a strong predictor of alcohol consumption in the first period after detoxification^44^. Furthermore, to support the validity of salivation as biomarker, it should be noted, that, higher mifepristone-induced cortisol level may induce acetylcholine production and, therefore, increased salivation (statistically reduced in both trials in the mifepristone, compared to the placebo, condition)^45^.

While this trial was developed based on our original preclinical^11^ and other translational^10^ literature related to mifepristone in AUD, subsequent preclinical studies in AUD model of rhesus monkeys showed that mifepristone decreased daily alcohol self-administration and the effect was mediated by mifepristone-induced increase in cortisol^18^. When we tested this hypothesis in our human study, first, we confirmed that cortisol increased after mifepristone administration, which is consistent with results in clinical setting^9,22^, human alcohol research^10^ and preclinical AUD models (alcohol dependent rats^10^ and monkeys^18^). However, we did not find that the effect of mifepristone on alcohol craving outcomes was mediated by increasing cortisol. Our results align with other clinical data showing that mifepristone’s effects on reducing psychotic symptoms were independent of the increased plasma cortisol and adrenocorticotropin hormone^9^. As this is a translational trial, we should evaluate further the discrepancy from the monkey study. First, the yohimbine administration was not done in monkeys. Also, it is possible that salivary cortisol (humans) is a more appropriate measure for adrenocortical function than blood (monkeys), as the salivary and serum total cortisol concentration have a non-linear relationship due to the rapid increase in salivary concentration once the serum cortisol-binding globulin is saturated^46^. Our clinical results, however, align with our preclinical data, which demonstrated that infusion of mifepristone directly into the amygdala suppressed yohimbine-induced reinstatement of alcohol-seeking, with corticosterone levels unaffected^11^. The involvement of the amygdala, rather than the effect on the negative feedback effect on the hypothalamus, was also later further supported by other translational AUD^10^ and psychiatric^9^ studies. Here, the mediation analysis suggests that the mifepristone effect in reducing yohimbine-induced alcohol craving is independent of the mifepristone-induced increase of cortisol level.

The second translational analysis was based on the work conducted in genetically selected msP rats, which demonstrated that mifepristone was less efficacious in reducing alcohol consumption compared to Wistar rats (rats trained to consume alcohol)^19^. This effect of mifepristone in msP rats is consistent with our clinical findings where mifepristone significantly reduced craving only in patients with *low* FHDA. In fact, the msP rat line is a well-characterized model of excessive voluntary alcohol drinking, and these lines are based on repeated generations of selective breeding for alcohol preference^47^. Therefore, mifepristone is less likely to be effective in patients who have supposedly a greater family contribution to the development of AUD^20,48^. It is also possible that mifepristone should be administered at a higher dose^10^ to block craving after repeated challenges. The dose response of mifepristone was also reported in a clinical study in patients with psychotic depression, where psychotic symptoms were reduced by mifepristone (1,200mg/day) and the effect was dependent on the blood level of mifepristone attained^9^.

This study provides exploratory and preliminary, yet promising, results on the potential role of GR antagonists in the treatment of AUD in a subtype population. There are possible confounding effects in this study by the placebo, as lower alcohol use was reported during placebo treatment for subjects with *low* FHDA. It is possible that the *low* FHDA group exhibited a differentially potent placebo effect, due to motivating conditions of the clinical trial^49^.

One of the major strengths of this study is the within-subject, cross-over design which provided the same set of participants acting as their own controls, increasing power and reducing variability^23^. The cortisol level also provided robust results for medication compliance, as it increases with mifepristone administration^22^. This is also the first time that yohimbine was paired to a cue-reactivity paradigm in a AUD population^12^, highlighting the translational efforts of this RCT between animal and human models^50^. The premise and the logistics of this work was paved by our preclinical study^11^ using the same medications paired to cue-induced reinstatement, stress-induced reinstatement and alcohol self-administration paradigms. The robustness of the study paradigm was also determined by our participants’ retention, regardless of the long washout period (3-week), required to allow the cortisol to reset at the basal level after the mifepristone administration.

A major limitation of this study was not having control placebo-conditions for yohimbine. Therefore, we could not determine if the effect of stress induction was due to the interaction between yohimbine combined to a cue-reactivity. Independently, however, both yohimbine and cue-reactivity are well-established and validated procedures for stress-induced alcohol craving and consumption^12^. Also, we did not collect mifepristone blood level in this work to ensure that clinical effects may be dependent on the blood level of mifepristone attained^9^. Finally, the low number of females enrolled in this study did not permit us to evaluate sex as a biological variable. In summary, this study provides important information on the safety of mifepristone as a medication to treat AUD. Our findings support the safety of mifepristone-alcohol combination in a human laboratory setting. The lack of serious or severe AEs, together with the low number of dropouts, indicate that mifepristone may be an acceptable medication in the treatment of AUD. Similarly, the safety data of this trial support the use of translational integration of yohimbine, combined to a cue-reactivity and alcohol self-administration protocol. In terms of efficacy, consistent with the earlier clinical work^10^, we found an effect in reducing alcohol craving, an important behavioral marker and now a diagnostic criterion in the DSM-5^51^. This translational trial fits with previous clinical studies that have utilized mifepristone in different psychiatry disorders, as mifepristone’s effects on reducing craving were independent of the increased plasma cortisol^9^. Future studies, with higher dose, are warranted to identify possible biomarkers of mifepristone’s response in patients with AUD and to best identify potential patients who will be responders or non-responders.

## Supporting information

Supplemental Material, Tables, Results

## Data Availability

All data produced in the present study are available upon reasonable request to the authors

## Acknowledgements

The authors thank Michael Brickley, Danielle Giovenco, Victoria Long, Zoe Brown (Brown University) and Karen Hafez (Corcept Therapeutics) for providing logistics support during the course of the trial. Also, we would like to thank Prabhjot Singh and Talia Vasaturo-Kolondier (Brown University) for providing help with the database.

## Authors Contributions

**CLH-K**: Lead for conceptualization, funding acquisition, investigation, methodology, project administration, analysis, writing-original draft, writing-review and editing. **CLH-K, PC, JCB, EGA** and **RMS** carried out the experiments and collected the experimental data. **CLH-K** and **MM** analyzed the data. **CLH-K, EGA, NC, RC, RS, JCB**, and **LL** provided substantial intellectual input for the interpretation of the results. All authors reviewed content and approved the final version of the manuscript.

