## Supplemental Material, Tables, Results for "Mifepristone as a pharmacological intervention for stress-Induced alcohol craving: a translational crossover randomized trial"

[1]Center for Alcohol and Addiction Studies, Brown University, Providence, RI, USA; [2]Department of Psychiatry and Human Behavior, Warren Alpert Medical School, Brown University, Providence, RI, USA; [3]Department of Behavioral and Social Sciences, School of Public Health, Brown University, Providence, RI, USA; [4]Carney Institute for Brain Science, Providence RI, Brown University; [5]School of Pharmacy, Pharmacology Unit, University of Camerino, Italy; [6]McLean Hospital, Belmont, MA, USA; [7]Division of Law, Ethics and Psychiatry, Department of Psychiatry, College of Physicians and Surgeons, Columbia University, New York City, NY, USA; [8]Yale Stress Center, Department of Psychiatry, Department of Neuroscience, Yale School of Medicine, Yale University, New Haven, CT; [9]Providence Veterans Affairs Medical Center, Providence, RI, USA; [10]Clinical Psychoneuroendocrinology and Neuropsychopharmacology Section, Translational Addiction Medicine Branch, NIDA IRP and NIAAA DICBR, Baltimore and Bethesda, MD, USA [11]Medication Development Program, National Institute on Drug Abuse Intramural Research Program, National Institutes of Health, Baltimore, MD, USA; [12]Division of Addiction Medicine, Department of Medicine, Johns Hopkins University School of Medicine, Baltimore, MD, USA; [13]Department of Neuroscience, Georgetown University Medical Center, Washington, DC, USA.

**[*]Corresponding Author:**
Carolina L. Haass-Koffler, PharmD, PhD
121 South Main Street
Providence, RI 02903


**Running Title**: Mifepristone for AUD
**RCT**: NCT02243709
**IND/FDA**: 121984, mifepristone and yohimbine

**Supplemental Materials: Methods S1**

*Participants*

*Inclusion criteria*: male or female individuals, between 21 and 65 years old (inclusive); current Diagnostic and Statistical Manual of Mental Disorders Text Revision four edition (DSM-IV-TR) diagnosis of alcohol dependence; participants had to meet criteria for moderate to heavy drinking (≥ 2 drinks/day for women and ≥ 3 drinks/day for men, during a 30-day period, within the 90 days prior to screening); participants had to be in good health as confirmed by medical history, physical examination, 12-lead electrocardiogram (ECG), and clinical laboratory tests; females had to be postmenopausal for at least one year, surgically sterile or using a barrier, non-hormonal birth control method (after a later IRB amendment); participants needed a BrAC=0.00 at each visit, be willing to take oral study medication and adhere to the study procedures.

*Exclusion criteria*: individuals seeking treatment for AUD; positive urine drug screen for substances (opioids, benzodiazepines, cocaine, methamphetamine, and tetrahydrocannabinol); diagnosed with a current substance use disorder, other than alcohol or nicotine; met criteria for a lifetime diagnosis of bipolar disorder, schizophrenia, or other psychotic disorders; active illness within the past six months of the screening visit that met the criteria for a diagnosis of Major Depressive Disorder (MDD) or Anxiety Disorder, or history of attempted suicide; clinically significant medical abnormalities: unstable hypertension, clinically significant abnormal ECG, bilirubin >150% of the upper normal limit (UNL), Alanine aminotransferase/Aspartate transaminase (ALT/AST) > 5 times the UNL, creatinine clearance ≤60dl/min; current use of psychotropic medications that may have an effect on alcohol consumption; current use of any medication involved in the metabolism of alcohol, such as aldehyde dehydrogenase (ALDH), alcohol dehydrogenase (ADH) and CYP2E1; current use of any medication (CYP3A4 inhibitor and substrate) that may interact with mifepristone; current use of any medication (CYP2D6 inhibitor and substrate) that may interact with yohimbine; history of seizure disorders; hypokalemia <3.5mEq/L; participated in any behavioral and/or pharmacological study within, minimum, the past 30 days; neuroendocrine disorders; taking corticosteroids; bleeding disorders; pre-existing QT prolongation on ECG (cut off 470ms female; 450ms male); history of porphyria; or not willing to engage in protected sex.

Note: Importantly, even though past clinical trials with mifepristone showed no increased depression[1,2], MDD and anxiety disorder were ascertained with SCID-IV, and depression and all anxiety disorders were assessed and excluded.

*Study procedures and measures*

Participants were recruited from the community in the RI and South MA area using advertising in social media and public transportation. Blinding was maintained by a research staff member not involved in the study. Randomization was performed using a computer allocation. The 3-week washout period was chosen to allow cortisol levels to return to baseline after mifepristone administration[3] and avoiding carryover effect, participants returned to the laboratory and received the crossover condition.

*Visit 1 (Screening)*: Following a breath analyzer to assess breath alcohol content (BrAC=0.00), participants signed a written informed consent. The screening assessments included clinical assessments: medical history, physical examination, vital signs, 12-lead ECG, blood and urine analysis, and psychiatric assessments: Structured Clinical Interview for DSM-IV-TR Axis I Disorders (SCID-IV)[4]; Hamilton anxiety rating scale (HAM-A)[5] and depression scale (HAM-D)[6]; Spielberg State and Trait Anxiety Inventory (STAI Y-1 and Y-2)[7]; Perceived Stress Scale (PSS)[8] and the Suicide Assessment Five-Step Evaluation and Triage (SAFE-T)[9]. Due to the distinct circadian rhythm of cortisol, all visits were scheduled approximately at the same time, in order to collect salivary samples to be measured both under basal "Rest" state (Visit 1) and induced "Stress" response (Visits 3 and 5) conditions. After a study physician reviewed and approved the medical history and clinical laboratory tests, the participants were scheduled for Visit 2.

*Visit 2 (Randomization)*: After controlling for a BrAC=0.00, other assessments included: TLFB, Life Events (LEC)[10], PTSD Checklist (PCL)[11], Brief Trauma Questionnaire (BTQ)[12], and study medication (mifepristone or placebo) was dispensed for seven days outpatient administration.

*Visits 3 and 5 (Alcohol Laboratory Session)*: The schematic outline of the laboratory paradigm procedures is depicted in **Figure 1** of the main manuscript. Participants were instructed not to consume alcohol for 24-hr, had to have a BrAC=0.00 and Clinical Institute Withdrawal Assessment for Alcohol-revised (CIWA-Ar)[13] score ≤10. A sample of saliva was collected and then a single oral dose of 32.4mg yohimbine was administered to participants. The cue reactivity began 30-min later in order to allow yohimbine to take effect[14], at which time a second sample of saliva was collected. The third saliva sample was collected after the cue reactivity. The cue reactivity was similar to previously published studies[15,16]. The water trial was included as a neutral control. Next, participants underwent two 3-min alcohol cue exposure trials that were identical to the water trial, except that the glass of water was replaced with their preferred alcoholic beverage. After every 3-min beverage exposure, participants rated their craving and urge to drink alcohol by completing the alcohol craving questionnaire (ACQ)[17], and the alcohol urge questionnaire (AUQ)[18]. The ACQ and AUQ both measure acute craving and they are both derived from the same construct[19], however, the AUQ was included as additional self-reported control construct, as it has shown high internal consistency, test-retest reliability, and has a strong correlation with alcohol dependent scale[17]. To extend from the self-reported assessments[20,21], we also included saliva (weight in cotton rolls, mg) monitored by passive droll at each trial of the cue reactivity. Systolic and diastolic blood pressure (mmHg) and heart rate (HR, beats/min) were monitored continuously. Following conclusion of the cue reactivity, participants underwent the alcohol self-administration procedure (priming alcohol drink and open bar). Participants received a priming dose of alcohol designed to raise blood alcohol levels to 0.03-0.05g/dl. Consistent with previous studies[22-26], we waited 40-min before presenting more alcohol. After the priming dose of alcohol, stimulant/sedative effects of alcohol were assessed using the Biphasic Alcohol Effects Scale (BAES)[22,27,28].

The “open bar” phase (alcohol self-administration, ASA) provided a total of eight standard drink unit (SDU) available and all may be consumed within 120-min, with four drinks presented during the first 60 minutes of the ASA, and then four new drinks during the other 60 minutes[22-24,29]. As an alternative reinforcer for not drinking, we provided $3 per each drink not consumed. Each beverage was calculated to raise the blood alcohol levels by 0.015g/dl. If the participant’s BrAC reached 0.1g/dl, the alcohol consumption ended. Participants waited until BrAC=0.00 and hemodynamics normalized before being discharged.

*Visits 4 and 6 (washout and follow-up)*: After a three-week washout period, participants returned to receive the opposite medication (placebo or mifepristone) for a week. Participants then returned for a 21-day follow-up for final assessments.

*Study drugs, dose justification and compliance*

Mifepristone does not require a titration and taper schedule. The dose for this study was based on the effect of mifepristone treatment (short and long duration)[30] in a clinical setting, which was shown to be safe and tolerable[31,32], including in patients with diagnosis of depression, anxiety, post-traumatic stress disorder (PTSD)[33] and AUD[34]. Compliance was monitored by pill count and, retrospectively, by salivary cortisol, as mifepristone increases the cortisol level by 10-fold compared to placebo[3].. The oral dose of yohimbine for this study was based on prior studies in which yohimbine was administered to examine neuroendocrine activation in human studies[33,35,36], and it was prepared and dispensed for each participant by a local compounding pharmacy. Ample data support the safe use of yohimbine at this target dose to induce craving in individuals with AUD[36,37].

*Statistical analysis*

Distributional characteristics of outcome measures were examined to evaluate similarity to the normal distribution, detailed descriptive analysis of demographics, substance use, and clinical characteristics were conducted. The sedation scale for the BAES, cortisol and amylase had a skewness and kurtosis in excess of two; consequently, an outlier analysis was performed and one outlier per group outside of ±3 interquartile range was treated as recommended[38]. Comparisons with these characteristics, in relation to enrolled versus completer status, were performed using *t*-tests to analyze continuous variables (e.g., age) and Chi square ($\chi^2$) for categorical variables (e.g., sex, race, smoking status). Attrition rates between the screening visit and follow-up visit were examined descriptively to assess for potential bias. In addition, a logistic regression was performed to test for possible bias due to period (A/B: placebo first, then mifepristone and B/A: mifepristone first, then placebo) or medication carryover (1: placebo and 2: mifepristone), as done in our prior cross-over trial[22].

*Primary outcomes:* safety and tolerability of oral administration of mifepristone was assessed after 7 days in outpatient setting, and when it was administered with yohimbine and alcohol during the laboratory paradigms.

These analyses included comparing the number of adverse events (AEs) between the mifepristone and placebo condition via a $\chi^2$ test. The results were presented using summary statistics such as number of subjects (*n*); mean (*M*); standard deviation (*SEM*) or frequency distributions (%). The safety and tolerability of mifepristone, compared to placebo, were also assessed by monitoring hemodynamic response, subjective effects of alcohol and pharmacokinetics parameters, using Generalized Estimating Equations (GEE)[39] with robust standard errors, and an unstructured correlation matrix. We conducted GEE with both laboratory paradigm procedures (time coded specifically for each analysis), medication (mifepristone/placebo) and visit (basal state and laboratory procedures) as within-subject factors. The model was specified to evaluate the effect of: 1) laboratory procedures (time/visit effect), 2) medication (mifepristone/placebo condition), and 3) the laboratory procedures by medication interaction. Hemodynamic response included: systolic (SBP) and diastolic (DBP) blood pressure, and heart rate (HR), with the laboratory paradigm procedures coded as time effect. Specifically, time was coded as $t_0$=yohimbine administration, $t_{30min}$=before and $t_{60min}$=after cue-reactivity. Alcohol pharmacokinetics parameters included: time to reach max concentration ($T_{max}$), max concentration ($C_{max}$), and area under the curve (AUC), calculated by $\int_{t=40}^{t=0}(BrAC)dx$ ($t_0$=time before and $t_{40min}$=40-min after prime alcohol administration), and were analyzed via data collected from the BrAC curve using confidence interval (*CI*) and interval estimate confidence, set at 95%. Subjective alcohol-related biobehavioral effects (stimulation/sedation) were measured by the Biphasic Alcohol Effects Scale (BAES)[28] on the alcohol biphasic curve. Data were collected from the breath alcohol content (BrAC) curve, time coded: $t_{10min}$=ascending and $t_{20min}$=descending limb.

*Secondary outcomes:* To evaluate the effect of stress (laboratory visit), compared to basal state (screening visit), on neuroendocrine variations in the absence of mifepristone, which increases the cortisol level by 10-fold[3], visit effect coded: $v_0$=basal, and $v_1$=laboratory; time coded: $t_0$=0 min (± laboratory procedures), $t_{30min}$=± before and $t_{60min}$=± after cue reactivity, with $t_0$=0 min (± yohimbine) inserted as a covariate.

*Alcohol consumption outcomes: A*lcohol consumption of the mifepristone condition, compared to placebo, was assessed in the bar laboratory after the alcohol priming dose and in the outpatient setting during the 7-day medication administration. In the outpatient setting, alcohol consumption was measured by self-report using the timeline follow back (TLFB) method, reported as heavy drinking days (HDD) and drinks per week (DPW), with time coded as $t_0$: baseline and $t_1$: after 7-day mifepristone/placebo administration.

*Neuroendocrine biomarker*: cortisol level collected by passive saliva drool, rather than blood, to avoid additional stress by needle.

All statistical analysis was performed after participants had their follow-up visit and the study database had been locked. All the statistical procedures were performed by IBM SPSS Statistics for Windows, version 27 with Macro MEMORE extension[9] (IBM Corp., Armonk, NY, USA) and GraphPad Prism (v.5) was used to generate figures (La Jolla, CA, USA). The effect sizes were calculated as $\eta^2$ for GLM and Cohen *d* for *t*-test. All statistical tests were two-tailed, and statistical significance was accepted if an alpha value $p$<.05 was obtained.

*Power*

This was a proof-of-concept trial to demonstrate the feasibility of the combined study design, the safety and tolerability of mifepristone and yohimbine while consuming alcohol, and the potential value of testing mifepristone in an appropriately-powered larger RCT. In selecting a target sample size, we balanced power considerations and feasibility given the translational nature of this trial. Because of the within-subjects design, power to test the effects of study drug is optimized for this modest sample size (originally $N$=20 then, after additional funding, increased to $N$=32). For the safety and tolerability outcomes (primary) adverse events difference was detected based on a judgement concerning the minimal effect which has clinical relevance in the management of patients. In a noninferiority trial, the exact sample size cannot be fixed in advance because it depends upon the chosen stopping guideline[40].

All statistical analysis was performed after participants had their follow-up visit and the study database locked. All the statistical procedures were performed by IBM SPSS Statistics for Windows, version 27 with Macro MEMORE extension (IBM Corp., Armonk, NY, USA) and GraphPad Prism (v.5) was used to generate figures (La Jolla, CA, USA). All statistical tests were two-tailed, and statistical significance was accepted if an $\alpha$ value $p<.05$ was obtained.

Protocol is accessible by contacting the PI: Prof. Carolina L. Haass-Koffler, PharmD, PhD

**Supplemental Materials: Results S1**

*Participants' characteristics, retention*

There was no difference in the attrition analysis conducted using period (placebo first, then mifepristone and mifepristone first, then placebo; $p>.05$) or medication (placebo and mifepristone; $p>.05$) as predictors. Five individuals withdrew from the study. In the mifepristone condition, one individual did not attend the first laboratory visit due to a family emergency ($n=1$, 3%). In the placebo condition, in the first laboratory session, one individual was not compliant with the laboratory procedures; one individual experienced a non-serious adverse event during the washout period, one participant was hospitalized for an event not related to the study procedures or study medication and for one individual we lost contact before attending the first laboratory procedures ($n=4$, 13%). For the second laboratory session, one individual in the placebo condition, was unable to complete the study due to COVID-19 in person restrictions ($n=1$, 3%), however the data in the naturalistic condition (no laboratory procedures) were completed with assessments collected remotely with an IRB-approved amendment[41].

*Safety and tolerability outcomes in outpatient setting*

During the 7-day administration of mifepristone or placebo in an outpatient setting, we did not observe serious AEs related to the study drugs or study procedure (**Table S2**). Mild to moderate non-serious AEs were reported by both study conditions throughout the trial, with no difference (mifepristone, $n=28$; placebo, $n=32$; $p's>.05$). In the placebo group, a total of 19 mild AEs were experienced by six participants and two moderate AEs were expressed by two participants. In the mifepristone condition, a total of 24 mild AEs were experienced by seven participants and six moderate AEs were experienced by two participants. The most frequently reported AEs in the placebo condition (with an incidence of ≥5%) were mild slowness, sleepiness, or fatigue in four participants (13%), mild difficulty with concentration or attention in two participants (6%), mild tremors in two participants (6%), and mild nausea in two participants (6%). The most frequently reported AEs in the mifepristone condition (with an incidence of ≥5%) were mild difficulty sleeping in two participants (7%), mild slowness, sleepiness, or fatigue in two participants (7%), mild restlessness in two participants (7%), mild nervousness or anxiety in 2 participants (7%), and mild irritability in two participants (7%). The reported adverse effects were mild (75% with placebo; 78% with mifepristone) or moderate (25% with placebo; 22% with mifepristone), and there were no severe adverse effects reported.

*HPA response*

The increase of cortisol in the mifepristone, compared to placebo, is aligned with results from preclinical AUD models (alcohol dependent rats[34] and monkeys[42]), human alcohol research[34] and other psychiatric (bipolar disorder, schizophrenia) research setting[3]. Here, we tested the increase of cortisol level as a mediator of alcohol craving, urge and cue-elicited salivary output after 7-day mifepristone administration.

The HPA response during the stress procedures was tested by comparing cortisol levels collected at the screening visit (basal) with the values collected during the laboratory procedures (yohimbine + cue-reactivity) at the same time points. Visit effect was coded as $v_0$=basal, and $v_1$=laboratory; time effect was coded as $t_{30min}$=± before cue reactivity and $t_{60min}$=± after cue reactivity, with baseline cortisol values before yohimbine administration ($t_0$) inserted as covariate. We found a significant main effect for visit (*b*=.072; *CI*=.51, .75; *p*=.002), such that higher cortisol was observed in the stress-induced visit compared to the screening visit (basal), no main effect for time (*p*>.05), but there was a visit by time interaction, such that a higher cortisol level occurred after the cue reactivity ($t_{60min}$: *b*=.141; *CI*=.44, .76, *p*<.001), suggesting an increase of HPA-axis response in the laboratory sessions (**Figure S1**).

*FHDA moderation analysis*

To test if FHDA moderated how participants responded to the laboratory procedures, the increase of the HPA response was confirmed when we compared the value of cortisol levels collected at the screening visit (basal) to the values collected during the laboratory visits (stress) in the placebo. When we compared the cortisol levels at the screening visit (basal) to the cortisol levels collected during the laboratory visits (stress), we found a significant main effect for FHDA (*b*=.41; *CI*=.10, 3.20; *p*=.002), such that higher cortisol was observed in the FHDA *low* group, and a visit by time interaction, such that there was a higher cortisol level during the laboratory procedures only in the FHDA *low* group (*b*=1.02; *CI*=2.90, 5.21, *p*=.002) (**Figure S2**), suggesting that in individuals with FHDA *low*, the HPA-axis responses were more activated during the laboratory procedures.

Moderation analysis of the drinking outcome in outpatient setting indicated no FHDA main effect (*p*>.05), but a FHDA by drug interaction showed a decrease in heavy drinking days for the *low* FHDA compared to the *high* FHDA (*p*=.014) from baseline; and a decrease in drinks per week for the *low* FHDA compared to the *high* FHDA in the placebo (*p*=.005) from baseline. There was no difference in the mifepristone group compared to placebo on both drinking outcomes (**Figure S3**).

**Supplemental Materials: Tables**

**Tables**

**Table S1** Sociodemographic and baseline clinical characteristics of the initial sample of participants who were enrolled and randomized in the study, expressed as n (%) or *M* ± (*SD*)

| | |
|---|---|
| Number (*N*) | 32 |
| Male, *n* (%) | 27 (84) |
| Hispanic, *n* (%) | 4 (13) |
| White, *n* (%) | 22 (69) |
| Age, *N* (*SD*) | 43 (12) |
| Marital status: married/relationship, *n* (%) | 11 (34) |
| Smoker, *n* (%) | 11 (34) |
| Cannabis, *n* (%) | 11 (34) |
| Age onset alcoholism (AOA) (*SD*) | 23 (8) |
| Baseline drinking days (DD) (*SD*) | 74 (17) |
| Baseline heavy drinking days (HDD) (*SD*) | 46 (30) |
| Baseline drinks per week (DPW) (*SD*) | 39 (26) |
| Alcohol dependence scale (ADS) (*SD*) | 8 (6) |
| Alcohol urge questionnaire (AUQ) (*SD*) | 23 (12) |
| Alcohol craving questionnaire (ACQ) | 43 (15) |
| Drinker inventory consequences (DrInc) (*SD*) | 32 (18) |
| Family history density alcoholism (FHDA) >66% (*SD*) | 17 (53) |
| Systolic blood pressure (SBP) (*SD*) | 126 (17) |
| Diastolic blood pressure (DBP) (*SD*) | 77 (9) |
| Heart rate (HR) (*SD*) | 74 (14) |
| Body Mass Index (BMI) (*SD*)* | 29 (11) |
| State-State Anxiety Inventory (STAI, state) (*SD*) | 31 (10) |
| State-Trait Anxiety Inventory (*STAI, trait*) (*SD*) | 34 (11) |
| Hamilton anxiety rating scale (HAMA) (*SD*) | 4 (5) |
| Hamilton depression rating scale (HAMD) (*SD*) | 3 (4) |
| Perceived stress scale (PSS) (*SD*) | 11 (6) |
| Post-traumatic stress disorder check list (PCL) *n* (%) | 24 (7) |
| PTSD criterion A (Brief Trauma questionnaire), *n* (%) | 18 (46) |
| PTSD criterion A (Life event checklist) *n* (%) | 26 (67) |

* Yohimbine dose was not adjusted for body weight as there was no difference in hemodynamic (SBP, DBP, HR, *p's*>.05), HPA axis (cortisol *p*>.05) response in individuals with different body mass index (BMI range 19-62 $kg/m^2$).

**Table S2 -** Non-serious adverse events [*number of AE and* percentage (%)]

| Side effect and severity | Placebo (*N*=31) | Mifepristone (*N*=27) | $X^2$ | *p* | *Cramer v-value* |
|---|---|---|---|---|---|
| Dizziness | | | | | |
| Mild | 0 | 0 | 1.126 | >.05 | 0.138 |
| Moderate | 0 | 1(4) | | | |
| Decrease in appetite | | | | | |
| Mild | 1(3) | 0 | 0.005 | >.05 | 0.010 |
| Moderate | 0 | 1(4) | | | |
| Increase in appetite | | | | | |
| Mild | 1(3) | 1(4) | 1.126 | >.05 | 0.138 |
| Moderate | 0 | 1(4) | | | |
| Difficulty sleeping | | | | | |
| Mild | 1(3) | 2(7) | 0.468 | >.05 | 0.089 |
| Moderate | 1(3) | 0 | | | |
| Slowness, sleepiness, or fatigue | | | | | |
| Mild | 4(13) | 2(7) | 0.534 | >.05 | 0.095 |
| Moderate | 1(3) | 0 | | | |
| Difficulty with coordination or balance | | | | | |
| Mild | 0 | 1(4) | 1.126 | >.05 | 0.138 |
| Moderate | 0 | 0 | | | |
| Difficulty with concentration or attention | | | | | |
| Mild | 2(6) | 1(4) | 0.253 | >.05 | 0.065 |
| Moderate | 0 | 0 | | | |
| Tingling in fingers or toes | | | | | |
| Mild | 0 | 1(4) | 1.126 | >.05 | 0.138 |
| Moderate | 0 | 0 | | | |
| Memory difficulties | | | | | |
| Mild | 0 | 1(4) | 1.126 | >.05 | 0.138 |
| Moderate | 0 | 0 | | | |
| Changes in taste | | | | | |
| Mild | 0 | 1(4) | 0.919 | >.05 | 0.125 |
| Moderate | 0 | 0 | | | |
| Tremor | | | | | |
| Mild | 2(6) | 1(4) | 0.919 | >.05 | 0.125 |
| Moderate | 0 | 0 | | | |
| Nausea | | | | | |
| Mild | 2(6) | 0 | 0.005 | >.05 | 0.010 |
| Moderate | 0 | 0 | | | |
| Vomiting | | | | | |
| Mild | 0 | 1(4) | 1.126 | >.05 | 0.138 |
| Moderate | 0 | 0 | | | |
| Headache | | | | | |
| Mild | 1(3) | 0 | 1.126 | >.05 | 0.138 |
| Moderate | 0 | 1(4) | | | |
| Restlessness | | | | | |
| Mild | 0 | 2(7) | 2.292 | >.05 | 0.197 |
| Moderate | 0 | 0 | | | |
| Nervousness or anxiety | | | | | |
| Mild | 1(3) | 2(7) | 0.468 | >.05 | 0.089 |
| Moderate | 0 | 0 | | | |

| **Side effect and severity** | **Placebo (*N*=31)** | **Mifepristone (*N*=27)** | $X^2$ | ***p*** | ***Cramer v-value*** |
|---|---|---|---|---|---|
| Irritability<br>Mild<br>Moderate | <br>1(3)<br>0 | <br>2(7)<br>0 | 0.468 | >.05 | 0.089 |
| Depression or other mood disturbance<br>Mild<br>Moderate | <br>1(3)<br>0 | <br>1(4)<br>0 | 0.919 | >.05 | 0.125 |
| Confusion<br>Mild<br>Moderate | <br>0<br>0 | <br>1(4)<br>0 | 1.126 | >.05 | 0.138 |
| Difficulty or inability to awaken from sleep<br>Mild<br>Moderate | 1(3)<br>0 | 1(4)<br>0 | 2.292 | >.05 | 0.197 |
| Difficulty breathing, wheezing, or cough<br>Mild<br>Moderate | <br>0<br>0 | <br>0<br>1(4) | 1.126 | >.05 | 0.138 |
| Swelling of throat or tongue<br>Mild<br>Moderate | <br>0<br>0 | <br>1(4)<br>0 | 1.126 | >.05 | 0.138 |
| Muscle aches<br>Mild<br>Moderate | <br>0<br>0 | <br>1(4)<br>0 | 1.126 | >.05 | 0.138 |
| Severe mood changes<br>Mild<br>Moderate | <br>1(3)<br>0 | <br>0<br>1(4) | 0.005 | >.05 | 0.010 |
| Other: | 0<br>0 | 1(4)<br>0 | 1.126 | >.05 | 0.138 |

**Supplemental Materials: Figures**

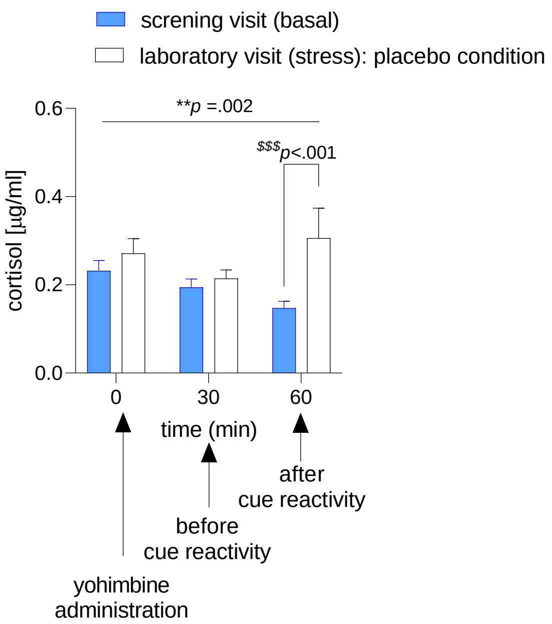


**Figure S1** - *Basal state (screening visit) compared to stress procedures (laboratory visit: yohimbine + cue-reactivity in the placebo condition only) on cortisol* (mifepristone, *n*=25; placebo, *n*=28). Cortisol: significant main effect for visit (*b*=.072; *CI*=.51, .75; *p*=.002), no main effect for time (*p*>.05); there was a visit by time interaction, after the cue reactivity ($t_{60min}$: *b*=.141; *CI*=.44, .76, *p*<.001). All data presented as mean±*SEM*. $^{*}p$ < .05 main effect; $^{\$}p$ < .05 interaction.

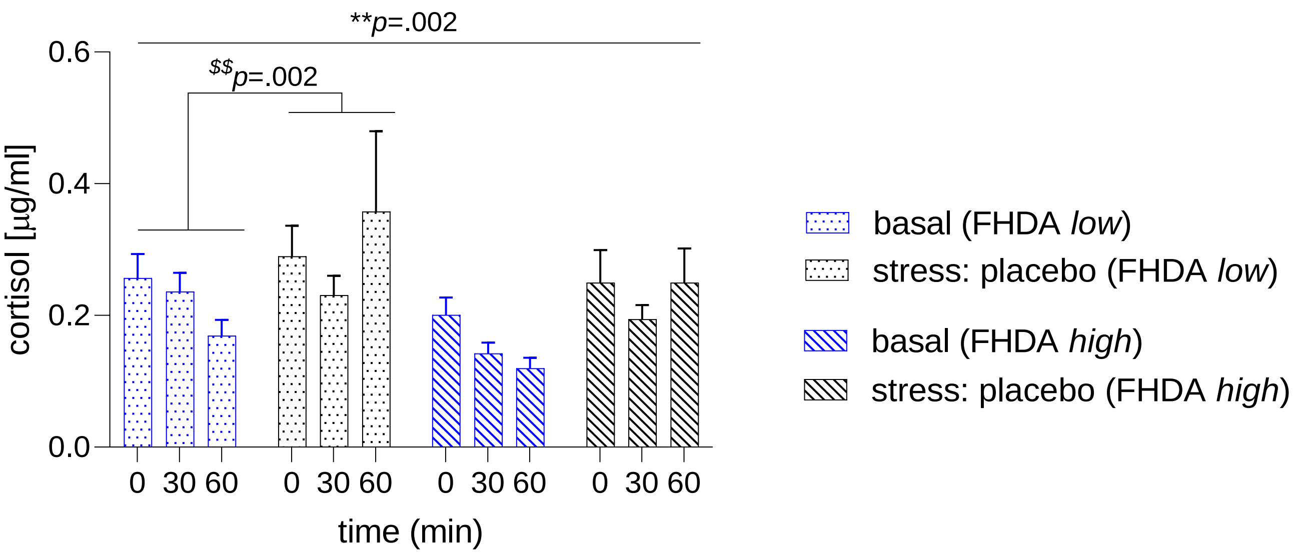


**Figure S2 -** *FHDA moderation analysis: cortisol and α-amylase control at basal visit* (mifepristone, *n*=25; placebo, *n*=28). cortisol: a significant main effect for FHDA (*b*=.41; *CI*=.10, 3.20; *p*=.002) and a visit by time interaction (*b*=1.02; *CI*=2.90, 5.21, *p*=.002). All data presented as mean±*SEM*. $^{*}p<.05$ main effect; $^{\$}p<.05$ interaction.

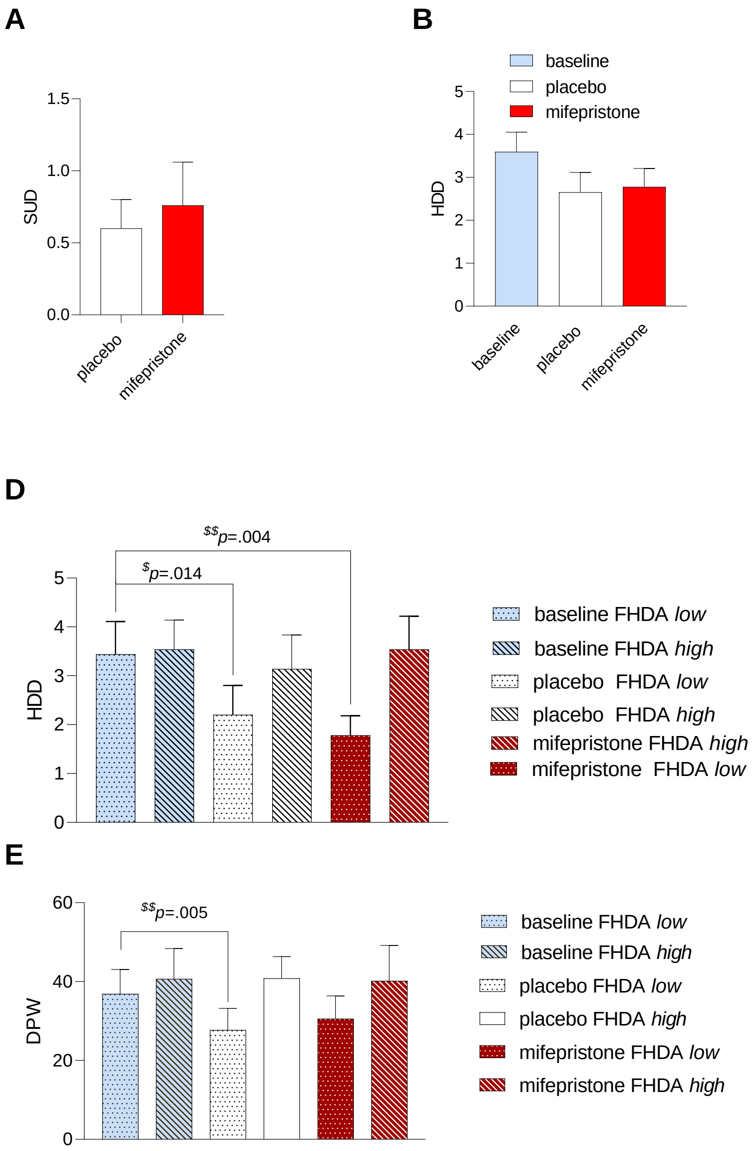


**Figure S3** - *The effect of mifepristone compared to placebo on alcohol consumption* (mifepristone, $n$=25; placebo, $n$=28)*.* **A**) There was no significant effect of mifepristone compared to placebo on alcohol consumption in the laboratory ($p>.05$). There was no significant effect of mifepristone compared to placebo on alcohol consumption in the outpatient setting ($p>.05$): **B**) Heavy Drinking Days (HDD) and **C**) Drinks Per Week (DPW). *FHDA moderator analysis:* **D** and **E**) There was no FHDA main effect ($p>.05$) and FHDA by medication interaction showed a decrease in HDD for the *low* FHDA compared to the *high* FHDA in the placebo ($p=.014$) and mifepristone ($p=.004$) group from baseline; and a decrease in DPW for the *low* FHDA compared to the *high* FHDA in the placebo ($p=.005$) from baseline. There was no difference in the mifepristone group compared to placebo on HDD and DPW. All data presented as mean±*SEM*, significance ($p<.05$). $^{*}p<.05$ main effect; $^{\$}p<.05$ interaction.

**Supplemental Materials: References**


1. Flores BH, Kenna H, Keller J, Solvason HB, Schatzberg AF. Clinical and biological effects of mifepristone treatment for psychotic depression. *Neuropsychopharmacology*. Mar 2006;31(3):628-36. doi:10.1038/sj.npp.1300884
2. DeBattista C, Belanoff J, Glass S, et al. Mifepristone versus placebo in the treatment of psychosis in patients with psychotic major depression. *Biol Psychiatry*. Dec 15 2006;60(12):1343-9. doi:10.1016/j.biopsych.2006.05.034
3. Gallagher P, Watson S, Elizabeth Dye C, Young AH, Nicol Ferrier I. Persistent effects of mifepristone (RU-486) on cortisol levels in bipolar disorder and schizophrenia. *J Psychiatr Res*. Oct 2008;42(12):1037-41. doi:10.1016/j.jpsychires.2007.12.005
4. First MB. Structured clinical interview for DSM-IV axis I disorders. *Biometrics Research Department*. 1997;
5. Hamilton M. Diagnosis and rating of anxiety. *Br J Psychiatry*. 1969;3(special issue):76-79.
6. HAMILTON M. A rating scale for depression. 1960;
7. Spielberg C. Manual for the state-trate anxiety inventory. Consulting Psychologists Press. Calif; 1970.
8. Cohen S, Kamarck T, Mermelstein R. Perceived stress scale. *Measuring stress: A guide for health and social scientists*. 1994;10(2):1-2.
9. Jacobs D. Suicide Assessment Five-Step Evaluation and Triage (SAFE-T). 2009.
10. Gray MJ, Litz BT, Hsu JL, Lombardo TW. Psychometric properties of the life events checklist. *Assessment*. 2004;11(4):330-341.
11. Weathers FW, Litz BT, Herman DS, Huska JA, Keane TM. The PTSD Checklist (PCL): Reliability, validity, and diagnostic utility. San Antonio, TX;; 1993:
12. Schnurr P, Vielhauer M, Weathers F, Findler M. Brief Trauma Questionnaire. 1999;
13. Sullivan JT, Sykora K, Schneiderman J, Naranjo CA, Sellers EM. Assessment of alcohol withdrawal: the revised clinical institute withdrawal assessment for alcohol scale (CIWA-Ar). *British journal of addiction*. 1989;84(11):1353-1357.
14. Owen JA, Nakatsu SL, Fenemore J, Condra M, Surridge DH, Morales A. The pharmacokinetics of yohimbine in man. *Eur J Clin Pharmacol*. 1987;32(6):577-82. doi:10.1007/BF02455991
15. Monti PM, Rohsenow DJ, Hutchison KE, et al. Naltrexone's effect on cue-elicited craving among alcoholics in treatment. *Alcohol Clin Exp Res*. Aug 1999;23(8):1386-94.
16. Rohsenow DJ, Niaura RS, Childress AR, Abrams DB, Monti PM. Cue reactivity in addictive behaviors: theoretical and treatment implications. *Int J Addict*. 1990;25(7A-8A):957-93. doi:10.3109/10826089109071030
17. Singleton E, Tiffany S, Henningfield J. Development and validation of a new questionnaire to assess craving for alcohol: problems of drug dependence. 1994:289.

### CONSORT 2010 checklist of information to include when reporting a randomised trial*

| Section/Topic | Item No | Checklist item | Reported on page No |
|---|---|---|---|
| **Title and abstract** | | | |
| | 1a | Identification as a randomised trial in the title | 1 |
| | 1b | Structured summary of trial design, methods, results, and conclusions (for specific guidance see CONSORT for abstracts) | 2 |
| **Introduction** | | | |
| Background and objectives | 2a | Scientific background and explanation of rationale | 3 |
| | 2b | Specific objectives or hypotheses | 4 |
| **Methods** | | | |
| Trial design | 3a | Description of trial design (such as parallel, factorial) including allocation ratio | 4 |
| | 3b | Important changes to methods after trial commencement (such as eligibility criteria), with reasons | 4 |
| Participants | 4a | Eligibility criteria for participants | 2, Methods S1 |
| | 4b | Settings and locations where the data were collected | 4 |
| Interventions | 5 | The interventions for each group with sufficient details to allow replication, including how and when they were actually administered | 3 |
| Outcomes | 6a | Completely defined pre-specified primary and secondary outcome measures, including how and when they were assessed | 5 |
| | 6b | Any changes to trial outcomes after the trial commenced, with reasons | 3 |
| Sample size | 7a | How sample size was determined | 3, Methods S1 |
| | 7b | When applicable, explanation of any interim analyses and stopping guidelines | NA |
| Randomisation: | | | |
| Sequence generation | 8a | Method used to generate the random allocation sequence | 4 |
| | 8b | Type of randomisation; details of any restriction (such as blocking and block size) | 4 |
| Allocation concealment mechanism | 9 | Mechanism used to implement the random allocation sequence (such as sequentially numbered containers), describing any steps taken to conceal the sequence until interventions were assigned | 4 |
| Implementation | 10 | Who generated the random allocation sequence, who enrolled participants, and who assigned participants to | 4, |

| | | | |
|---|---|---|---|
| | | interventions | Methods S1 |
| Blinding | 11a | If done, who was blinded after assignment to interventions (for example, participants, care providers, those assessing outcomes) and how | 4 |
| | 11b | If relevant, description of the similarity of interventions | NA |
| Statistical methods | 12a | Statistical methods used to compare groups for primary and secondary outcomes | 4 |
| | 12b | Methods for additional analyses, such as subgroup analyses and adjusted analyses | 5 |
| **Results** | | | |
| Participant flow (a diagram is strongly recommended) | 13a | For each group, the numbers of participants who were randomly assigned, received intended treatment, and were analysed for the primary outcome | 5 |
| | 13b | For each group, losses and exclusions after randomisation, together with reasons | 5 |
| Recruitment | 14a | Dates defining the periods of recruitment and follow-up | 5 |
| | 14b | Why the trial ended or was stopped | NA |
| Baseline data | 15 | A table showing baseline demographic and clinical characteristics for each group | 9 Results S1 |
| Numbers analysed | 16 | For each group, number of participants (denominator) included in each analysis and whether the analysis was by original assigned groups | 5 |
| Outcomes and estimation | 17a | For each primary and secondary outcome, results for each group, and the estimated effect size and its precision (such as 95% confidence interval) | 5 |
| | 17b | For binary outcomes, presentation of both absolute and relative effect sizes is recommended | 5 |
| Ancillary analyses | 18 | Results of any other analyses performed, including subgroup analyses and adjusted analyses, distinguishing pre-specified from exploratory | 8 |
| Harms | 19 | All important harms or unintended effects in each group (for specific guidance see CONSORT for harms) | NA |
| **Discussion** | | | |
| Limitations | 20 | Trial limitations, addressing sources of potential bias, imprecision, and, if relevant, multiplicity of analyses | 11 |
| Generalisability | 21 | Generalisability (external validity, applicability) of the trial findings | 11 |
| Interpretation | 22 | Interpretation consistent with results, balancing benefits and harms, and considering other relevant evidence | 11 |
| **Other information** | | | |
| Registration | 23 | Registration number and name of trial registry | 2 |
| Protocol | 24 | Where the full trial protocol can be accessed, if available | 6 Methods S1 |
| Funding | 25 | Sources of funding and other support (such as supply of drugs), role of funders | |

*We strongly recommend reading this statement in conjunction with the CONSORT 2010 Explanation and Elaboration for important clarifications on all the items. If relevant, we also recommend reading CONSORT extensions for cluster randomised trials, non-inferiority and equivalence trials, non-pharmacological treatments, herbal interventions, and pragmatic trials. Additional extensions are forthcoming: for those and for up to date references relevant to this checklist, see www.consort-statement.org.
